# Effect of the WHO Labour Care Guide Implementation on Maternal and Newborn Outcomes in Selected Referral Hospitals of Zambia: A Difference-in-Differences Analysis

**DOI:** 10.64898/2026.10.06.26364926

**Authors:** Makasa Musonda, Mangala Benedictus, Kachali Lambwe, Vwalika Bellington

## Abstract

**Background:** Despite being originally developed to support labour monitoring in low-resource and rural settings, the partograph has remained the standard tool for labour management and continues to be used by obstetricians and midwives across diverse settings worldwide. However, inappropriate use and non-use of the partograph persist and have been associated with increased odds of adverse perinatal outcomes, including perinatal death. To address these limitations, the World Health Organization introduced the Labour Care Guide in 2020 as a comprehensive tool to standardise and improve the quality of care during childbirth. Henceforth, some countries such as Rwanda and Uganda have since implemented its use. This study aimed to assess the effect of WHO Labour Care Guide implementation, compared with the traditional partograph, on selected maternal and perinatal outcomes across participating facilities.

**Methods:** This was a quasi-experimental implementation study conducted in selected referral hospitals in Zambia, comparing maternal and perinatal outcomes before and after implementation of the WHO Labour Care Guide. Facilities were grouped into implementation and comparison sites. Routine HMIS/DHIS2 data were used to assess selected outcomes across the pre- and post-implementation periods. Descriptive statistics were used to summarise outcome trends, while a difference-in-differences analytical approach was applied using Stata version 18 to compare changes over time between implementation and comparison sites.

**Results:** The analysis included 12 hospitals, 192 facility-month observations, 89,122 total births, and 87,004 live births. Overall, the stillbirth rate was 23.8 per 1,000 total births, birth asphyxia rate was 40.1 per 1,000 live births, caesarean section rate was 34.6 per 100 live births, low-birthweight rate was 168.6 per 1,000 live births, and maternal mortality ratio was 313.8 per 100,000 live births. The implementation sites reporting higher birth asphyxia rates (42.7 versus 38.0 per 1,000 live births). Difference-in-differences analysis showed no statistically significant effect of LCG implementation on stillbirths, IRR 1.07 (95% CI: 0.69–1.67, p=0.752); fresh stillbirths, IRR 1.23 (95% CI: 0.74–2.05, p=0.431); birth asphyxia, IRR 0.65 (95% CI: 0.34–1.27, p=0.207); or caesarean section, IRR 1.02 (95% CI: 0.80–1.30, p=0.892).

**Conclusion:** Implementation of the WHO Labour Care Guide was not associated with statistically significant changes in stillbirths, fresh stillbirths, birth asphyxia, or caesarean section rates during the study period. Despite the higher birth asphyxia rates in the implementation sites the difference remained statistically not significant. Scale-up of the LCG should be accompanied by sustained training, mentorship, audit and feedback, and strengthening of emergency obstetric and newborn care systems.

## Introduction

Childbirth is a momentous occasion that requires efficacious and evidence-based care to ensure positive pregnancy experience for the mother and neonate. To ensure this, most guideline for normal human labour progress are derived from Friedman’s clinical observations of women in labour (1). The partograph is a tool that provides a pictorial presentation of labour and provides a good overview of the labour progress (2). The concept of the partogram was introduced in 1954 by an American obstetrician and gynaecologist and labour researcher by graphically plotting cervical dilatation against time, the sigmoid curve, and divided the first stage of labour into latent and active phases. The latter was further divided into acceleration, maximum slope and deceleration (3, 4). Building on Friedman’s graphic labour curve, Robert Hugh Philpott a professor of obstetrics and gynaecology and Dr William Malcom Castle, University of Rhodesia (now University of Zimbabwe), Salisbury (Harare, Zimbabwe) developed a modified cervicograph/partograph for use in labour management and introduced the alert line and action line (5). In 1987, the World Health Organisation (WHO) launched the safe motherhood initiative to reduce maternal and perinatal morbidity and mortality by improving the quality and safety of maternity care. As part of the endeavours to strengthen the monitoring and management of labour, WHO subsequently developed and revised the partograph, publishing three different versions over time (1). WHO modified the partograph in 2000 excluded the latent phase and active phase commenced at 4cm cervical dilatation (6).

Despite the initial intent of the partograph having been developed for use in the developing world and rural settings, the partograph has remained the standard to labour management today and is used by obstetricians and midwives worldwide and in diverse settings (7). Even though its application has been extensive to practice, the partograph use has been incorrect in many settings, although even when used correctly, birth attendants have reported gaps in initiating proper labour management (8). Inappropriate and even non-use of the partograph still exist in some settings and is associated with significantly increased odds of adverse perinatal outcomes including death (9, 10). Zhang, Landy (11) provided important evidence that challenged the traditional Friedman labour curve and the assumptions underpinning the partograph. Using contemporary labour curve analyses, they demonstrated that labour in women with normal neonatal outcomes progressed more slowly than previously described by Friedman, particularly in the early active phase. Specifically, cervical dilatation between 4 cm and 6 cm was substantially slower than traditionally assumed, with labour taking more than 6 hours to progress from 4 to 5 cm and more than 3 hours from 5 to 6 cm in otherwise normal labour. Their findings further suggested that the active phase of labour begins closer to 6 cm rather than 4 cm, thereby questioning the appropriateness of applying traditional partograph thresholds too early in labour. This culminated in a major shift in obstetric practice, reflected in the American College of Obstetricians and Gynaecologists/ Society for Maternal-Foetal Medicine (ACOG/SMFM) 2014 consensus statement, Safe Prevention of the Primary Caesarean Delivery, which incorporated contemporary evidence into clinical guidance by recommending that standards of active labour should not be applied before 6 cm of cervical dilatation. To mitigate this, in 2020 WHO developed a comprehensive Labour Care Guide (LCG) to standardise and make better the quality of care received during childbirth. The LCG was intended to ensure individualised or women centred care so as to maximise the chance of positive pregnancy and childbirth experience (12). The development of the LCG was as a result of questions on how and whether the design of the partograph is suitable, optimum, and or effective enough suggesting migration from the partograph.

The WHO LCG, often described as the “future partograph,” was recommended for adoption by countries as a more person-centred and evidence-based approach to monitoring labour and childbirth. During the joint conference of the Zimbabwe Society of Obstetricians and Gynaecologists (ZSOG) and the East, Central and Southern Africa College of Obstetricians and Gynaecologists (ECSACOG), held in Victoria Falls from 12 to 15 October 2022, WHO formally encouraged member countries in the East, Central and Southern Africa region to adopt and implement the LCG. Although some countries have since introduced the LCG into routine maternity care, uptake across the region has generally been slow and uneven. Against this background, a pilot implementation study was undertaken in selected hospitals in the Copperbelt and Lusaka Provinces of Zambia to evaluate the effectiveness of the WHO LCG in routine maternity care. Accordingly, this study aimed to assess the effect of the WHO LCG implementation compared to the traditional partograph on selected maternal and perinatal outcomes across the participating facilities.

## Methods and Materials

### Study Design

We employed a quasi-experimental controlled before-and-after design to assess the implementation of the WHO LCG in selected hospitals. A Difference-in-Differences (DiD) analytical approach was applied to compare changes in maternal and newborn outcomes between implementation and control sites before and after introduction of the LCG. This method enabled us to estimate the effect of the LCG while accounting for underlying secular trends that may have affected both groups.

### Study Setting

The study was undertaken in twelve public-sector maternity facilities in Lusaka and Copperbelt Provinces of Zambia. Six facilities implemented the WHO LCG intervention (Chilenje First Level Hospital, Kafue General Hospital, Levy Mwanawasa University Teaching Hospital, Ndola Teaching Hospital, Ronald Ross Hospital, and Roan Antelope Hospital), while six facilities served as controls and continued standard intrapartum care practices (Women and Newborn Hospital, Chongwe District Hospital, Chipata First Level Hospital, Chifubu District Hospital, Kitwe Teaching Hospital, and Nchanga North Referral Hospital). These facilities represent primary-, secondary-, and tertiary-level maternity care institutions and collectively manage a substantial proportion of facility-based deliveries within their respective catchment areas. These sites providecomprehensive maternity services and contribute routinely to district and provincial and newborn health reporting systems.

### Intervention

The intervention consisted of the implementation of the WHO LCG. The implementation activities included health care worker orientation and training, introduction of the LCG documentation tools, supportive supervision, and routine monitoring of implementation fidelity. The control facilities continued standard labour monitoring practices with the partograph without knowing of the ongoing implementation of the LCG in selected sites.

### Study Period

The time framework covered for evaluation was 16-months from January 2025 to April 2026. For analytical purposes, the study was divided into two equal phases: an eight-month pre-implementation period from January 2025 to August 2025 (8 months), and an eight-month post-Implementation period: September 2025 to April 2026 (8 months).

### Study Population

The population for the study was comprised of women delivering in the participating hospitals during the time of the study and whose outcomes were recorded in routine maternity ward registers. The unit of analysis was the facility-month and not the patients individually. As a result, the analytical dataset consisted of aggregated monthly counts of maternal and perinatal outcomes for each facility. A total of 192 facility-month observations was included in the analysis, consisting of 96 observations from intervention facilities and 96 observations from control facilities.

### Data Sources

Monthly aggregated maternity and newborn health data were recorded from routine facility reporting systems. Data were compiled into a standardized electronic database and verified against source reports before analysis. The extracted indicator included institutional deliveries, live births, stillbirths, fresh stillbirths, macerated stillbirths, caesarean deliveries, birth asphyxia cases,low birth weight (<2500g), and maternal deaths.

The primary outcomes for the evaluation were stillbirth rate, fresh stillbirth rate, and birth asphyxia rate. Secondary outcomes included caesarean section rate, macerated stillbirth rate, and low birth weight rate. Total births were calculated as the sum of live births and stillbirths. Stillbirth, fresh stillbirth, and macerated stillbirth rates ere expressed per 1,000 total births. Birth asphyxia and low birth weight rates were expressed per 1,000 live births, while caesarean section rate was expressed per 100 live births. Maternal mortality ratios were expressed as percentages of live births. Maternal mortality ratios were expressed per 100,000 live births.

### Data Management

Data were entered into Microsoft Excel and imported into Stata version 18 (StataCorp LCC, College Station, Texas, USA) for analysis.

The original data was transformed from wide format to a facility-month information from the District Health Information Management System 2 (DHIMS2). Variables identifying implementation status, study period, and outcome indicators were generated. Data cleaning procedures included consistency checks, verification of denominators, identification of outliers, and assessment of missing values.

### Statistical Analysis

Descriptive statistical analyses were used to summarise facility characteristics and outcome measures by study arm and implementation period. Aggregate counts and rates were calculated using appropriate denominators. Graphical trend analyses were also conducted to examine monthly changes in selected outcome rates between intervention and control facilities for caesarean section and birth asphyxia rates, with vertical reference line indicating the start of the LCG implementation in September 2025.

To estimate the effect of the LCG implementation, Poisson regression models with robust standard errors were fitted for each outcome. A DiD specification was used by including an interaction term between implementation status and study period. Results were reported as incidence rate ratios (IRRs) with corresponding 95% confidence intervals (CI) and p-values. Statistical significance was assessed at α = 0.05.

## Results

Table 1 shows the evaluation which included 12 hospitals, with six control sites and six implementation sites. Each arm contributed 96 facility-month observations, giving a total of 192 facility-month observations over a 16-month period from January 2025 to April 2026. The pre-implementation period covered September 2025 to April 2026. Control sites continued routine labour monitoring with the partograph, whereas implementation sites introduced the WHO LCG.

**Table 1.** Study Characteristics and Aggregate Birth Outcomes by Study Arm.

| Characteristic | Control Sites | Implementation Sites | Total |
| --- | --- | --- | --- |
| Number of hospitals | 6 | 6 | 12 |
| Facility-month observations | 96 | 96 | 192 |
| Observation period | Jan 2025-Apr 2026 | Jan 2025-Apr 2026 | 16 months |
| Pre-implementation period | Jan-Aug 2025 | Jan-Aug 2025 | 8 months |
| Post-implementation period | Sep 2025-Apr 2026 | Sep 2025-Apr 2026 | 8 months |

| Intervention status | Routine labour<br>monitoring with<br>Partograph | WHO Labour Care<br>Guide implementation | - |
| --- | --- | --- | --- |

In table 2 across the 12 participating hospitals, a total of 87,004 live births and 89,122 total births were recorded during the study period. Control sites accounted for 48,873 live births and 50,187 total births, while implementation sites contributed 38,131 live births and 38,935 total births. Overall, 2,118 stillbirths were reported, including 669 fresh stillbirths and 1,449 macerated stillbirths. A total of 30, 120 caesarean sections, 3,486 cases of birth asphyxia, and 14,667 lbw births were recorded across all facilities, Additionally, 273 maternal deaths occurred during the study period, of which 182 were reported in control sites and 91 implementation sites.

**Table 2.** Aggregate births and outcomes.

| Characteristic | Control<br>Sites | Implementation<br>Sites | Total |
| --- | --- | --- | --- |
| Live births | 48,873 | 38,131 | 87,004 |
| Total births (live births + stillbirths) | 50,187 | 38,935 | 89,122 |
| Stillbirths | 1,314 | 804 | 2,118 |
| Fresh stillbirths | 348 | 321 | 669 |
| Macerated stillbirths | 966 | 483 | 1,449 |
| Caesarean sections | 16,505 | 13,615 | 30,120 |
| Maternal deaths | 182 | 91 | 273 |
| Birth asphyxia cases | 1,856 | 1,630 | 3,486 |
| Low birthweight births (<2500g) | 8,899 | 5,768 | 14,667 |

Table 3 presents the overall outcome rates across the study facilities. The overall stillbirth rate was 23.8 per 1,000 total births, with a higher rate observed in control facilities compared with implementation facilities (26.2 versus 20.6 per 1,000 live births). In contrast, implementation facilities recorded a slightly higher fresh stillbirth rate than control facilities (8.2 versus 6.9 per 1,000 total births).

**Table 3.**
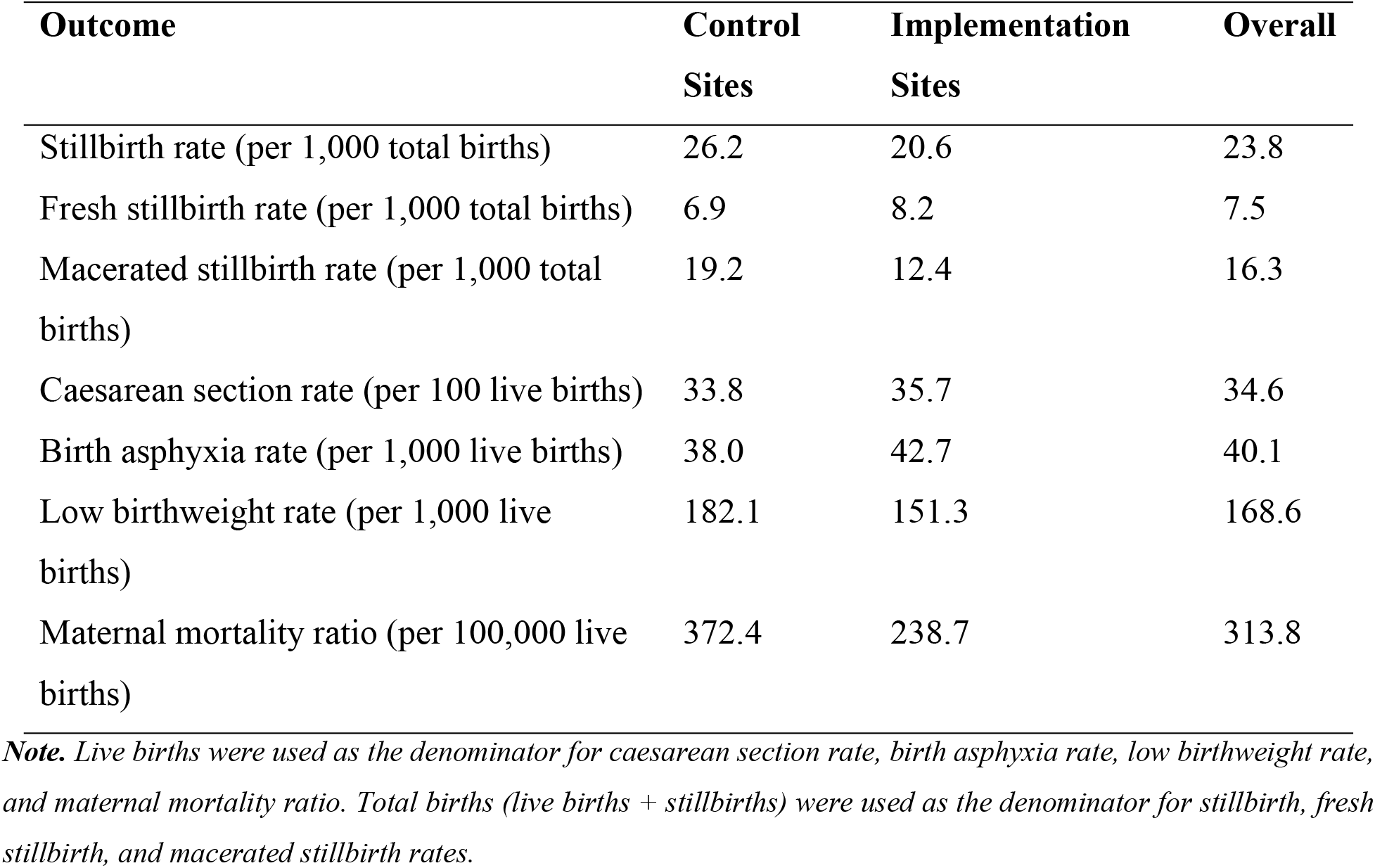
Derived rates.

| Outcome | Control<br>Sites | Implementation<br>Sites | Overall |
| --- | --- | --- | --- |
| Stillbirth rate (per 1,000 total births) | 26.2 | 20.6 | 23.8 |
| Fresh stillbirth rate (per 1,000 total births) | 6.9 | 8.2 | 7.5 |
| Macerated stillbirth rate (per 1,000 total<br>births) | 19.2 | 12.4 | 16.3 |
| Caesarean section rate (per 100 live births) | 33.8 | 35.7 | 34.6 |
| Birth asphyxia rate (per 1,000 live births) | 38.0 | 42.7 | 40.1 |
| Low birthweight rate (per 1,000 live<br>births) | 182.1 | 151.3 | 168.6 |
| Maternal mortality ratio (per 100,000 live<br>births) | 372.4 | 238.7 | 313.8 |

***Note.** Live births were used as the denominator for caesarean section rate, birth asphyxia rate, low birthweight rate,* *and maternal mortality ratio. Total births (live births + stillbirths) were used as the denominator for stillbirth, fresh* *stillbirth, and macerated stillbirth rates.*

The overall caesarean section rate was 34.6 per 1,000 live births and was marginally higher in implementation facilities than in control facilities (35.7% versus 33.8%). Birth asphyxia occurred at a rate of 40.1 per 1,000 live births overall, with implementation sites reporting higher rates (42.7 versus 38.0 per 1,000 live births), even though the absolute number was higher in the control sites. Lbw occurred at a rate of 168.6 per 1,000 live births overall and were more common in control facilities than implementation facilities (182.1 versus 151.3 per 1,000 live births). The overall maternal mortality ratio was 313.8 per 1,000 live births, with control facilities recording a substantially higher ratio than implementation facilities (372.4 versus 238.7 per 100,000 live births). These differences represent crude outcome rates and provide baseline comparison between the two study arms prior to formal assessment of intervention effects using DiD regression models.

Table 4 Difference-in-differences Poisson regression models were fitted to assess the effect of WHO Labour Care Guide implementation on maternal and perinatal outcomes. The interaction term between implementation status and study period represented the estimated intervention effect. No statistically significant intervention effect was observed for stillbirths (IRR = 1.07, 95% CI: 0.69–1.67, p = 0.752), fresh stillbirths (IRR = 1.23, 95% CI: 0.74–2.05, p = 0.431), birth asphyxia (IRR = 0.65, 95% CI: 0.34–1.27, p = 0.207), or caesarean section rates (IRR = 1.02, 95% CI: 0.80– 1.30, p = 0.892).

**Table 4.** Difference-in-Differences Poisson Regression Analysis of WHO Labour Care Guide Implementation on Maternal and Perinatal Outcomes.

| Outcome | IRR | 95% CI | p-value |
| --- | --- | --- | --- |
| Stillbirths | 1.07 | 0.69–1.67 | 0.752 |
| Fresh stillbirths | 1.23 | 0.74–2.05 | 0.431 |
| Birth asphyxia | 0.65 | 0.34–1.27 | 0.207 |
| Caesarean section | 1.02 | 0.80–1.30 | 0.892 |

**Figure 1.**
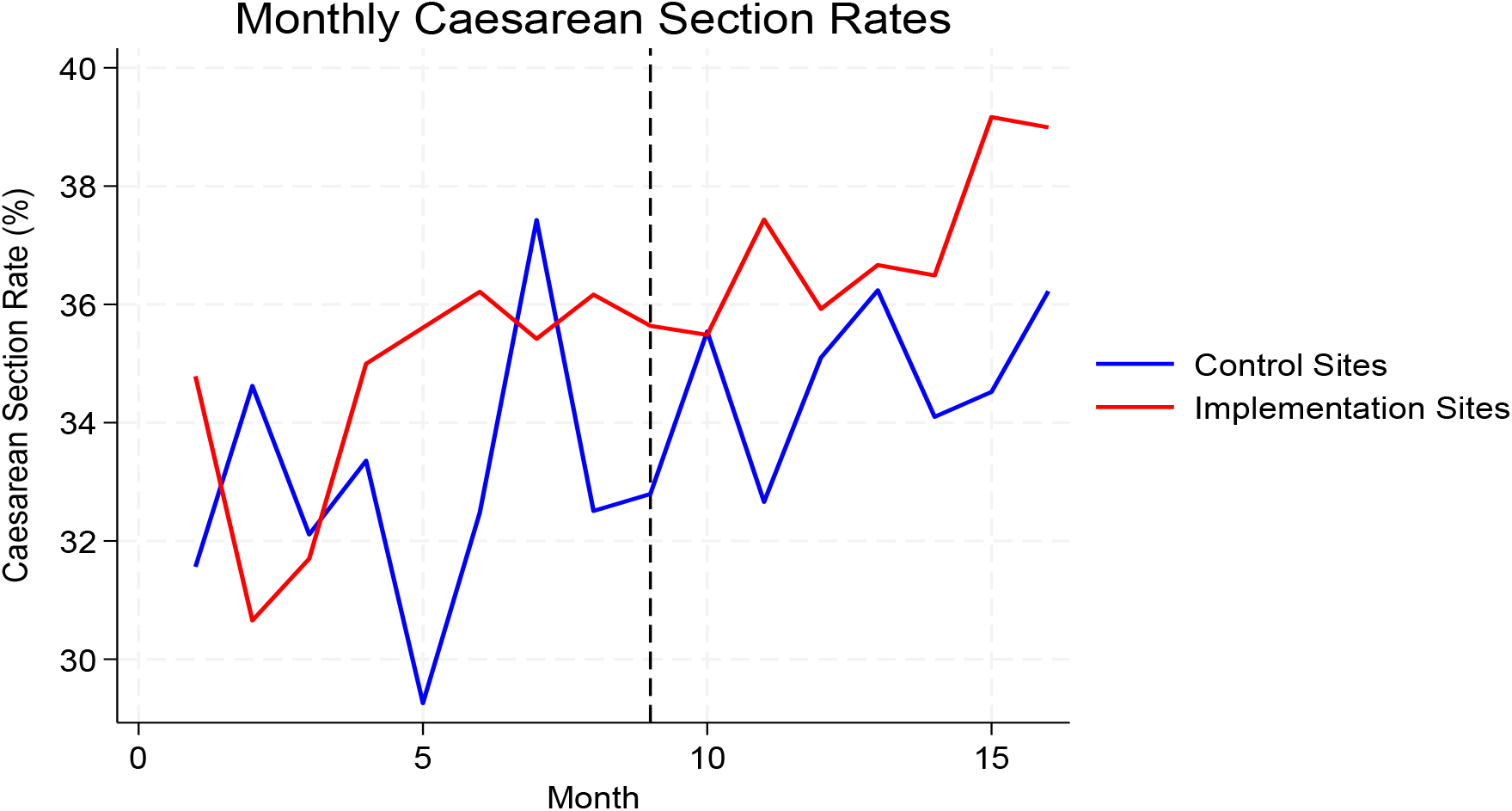
Monthly Caesarean Section Rates in Control and Implementation Sites, January 2025–April 2026. The dashed vertical line indicates introduction of the WHO Labour Care Guide in September 2025. Caesarean section rates remained relatively stable throughout the study period in both implementation and control sites, with no clear divergence following implementation. These findings are consistent with the difference-in-differences Poisson regression analysis, which found no statistically significant intervention effect on caesarean section rates (IRR = 1.02, 95% CI: 0.80–1.30; p = 0.892).

**Figure 2.**
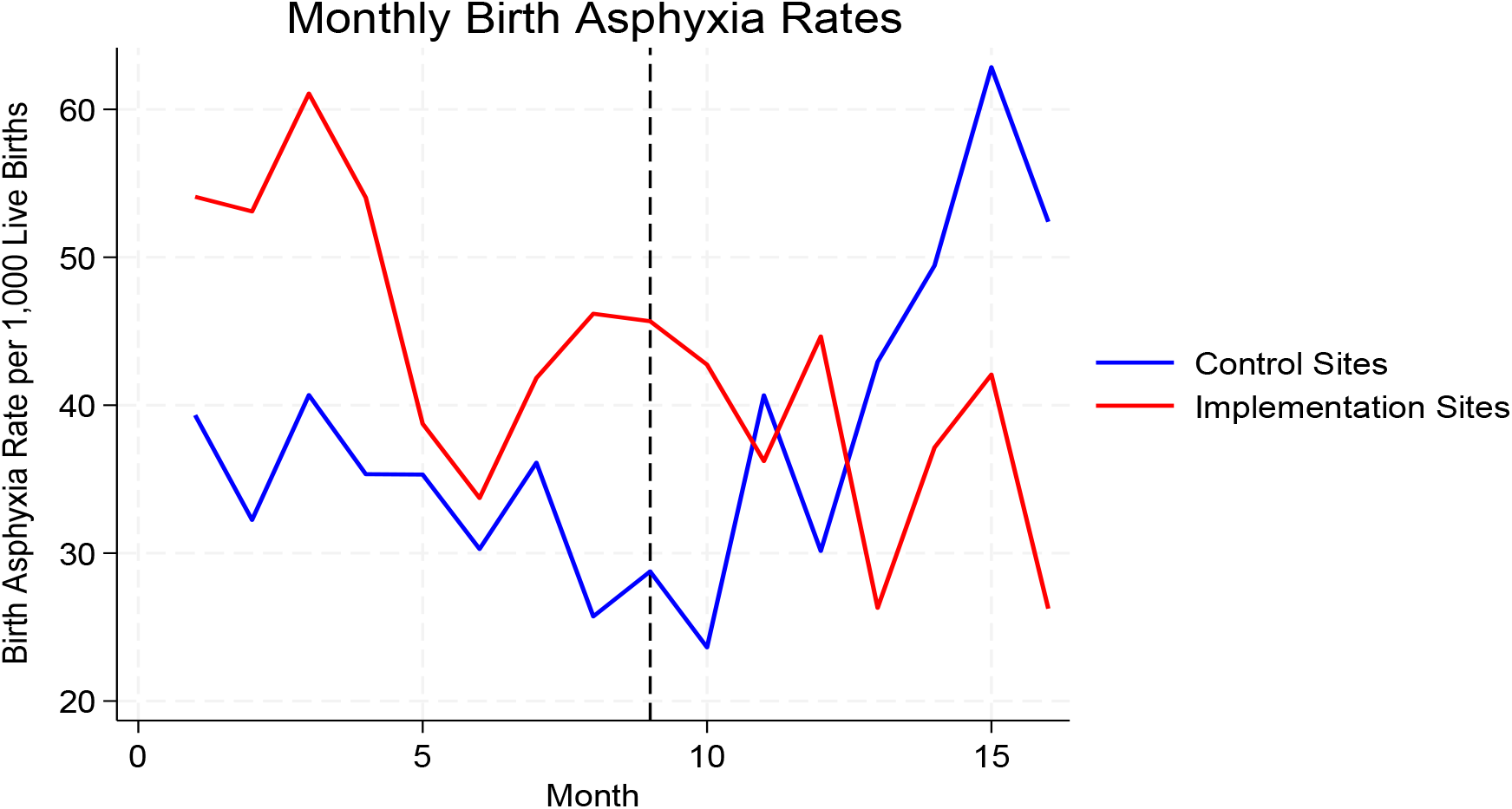
Monthly Birth Asphyxia Rates in Control and Implementation Sites, January 2025–April 2026. Figure 2 shows that birth asphyxia rate showed marked month-to-month variation in both study arms. Prior to LCG implementation, rates were generally higher in implementation sites than control sites. After implementation, rates in implementation site showed on overall downward trend, while control sites show greater variability and an increase in later post-implementation months. This visual pattern suggested a possible reduction in birth asphyxia following LCG implementation, although DiD regression do not show statistically significant intervention effect. Monthly birth-asphyxia rates varied considerably throughout the study period. In the post-implementation period, the implementation sites showed fluctuating rates with declines in some months, while the control sites demonstrated an overall upward trend, particularly during the later months. The apparent divergence between the two groups was further evaluated using DiD analysis.

## Discussion

This quasi-experimental evaluation assessed the effect of the WHO LCG implementation compared to the traditional partograph on maternal and newborn outcomes across participating hospitals of the Copperbelt and Lusaka Provinces of Zambia. Overall, the DiD analysis showed that the intervention was not associated with statistically significant improvements in the evaluated outcomes. Although implementation sites demonstrated a 35% relative reduction in birth asphyxia compared with control sites in the adjusted model, this effect did not reach statistical significance. Taken together, these findings suggest that while the LCG may have shown potential to improve selected intrapartum outcomes, particularly birth asphyxia, its introduction over the study period was not associated with clear measurable reductions in the major maternal and perinatal outcomes examined.

While most previous studies show that the WHO LCG is associated with significant reductions in caesarean section rates compared to the control group that used the traditional partograph (13, 14). On the contrary, our study did not demonstrate significant reduction in caesarean section rates in the WHO LCG implementation sites. This is in keeping with another study by Misra, Koushar (15) that reported comparable caesarean section rates and instrumental deliveries between LCG and conventional partograph monitored groups. On the other hand, the LCG use has also been reported to be associated with reduced emergency caesarean sections as it promotes individualised labour assessment rather than fixed time thresholds, while Mugyenyi, Tumuhimbise (16) also reported a decline in emergency caesarean deliveries in rural settings following LCG implementation (17). The findings collectively are suggestive that the LCG implementation facilitates physiological labour progression and reduces unnecessary surgical interventions.

The key finding of this study is that stillbirth rates did not differ significantly either following LCG implementation. The adjusted DiD model showed no significant intervention effect on stillbirths overall or on fresh stillbirth specifically. This consistent with previous studies that also reported no significant differences observed in neonatal outcomes between the LCG monitored group and the conventional partograph group (17, 18). This is an important finding given that the LCG was introduced to improve labour monitoring, support timely recognition of abnormal labour, and strengthen decision-making during intrapartum care. One plausible reason is that stillbirths are influenced by a broad range of factors that extend beyond intrapartum care alone. Evidence from Zambian studies conducted in settings similar to the present one suggests that stillbirth and perinatal mortality are strongly influenced by both maternal and health-system factors. Reported predictors include late antenatal care booking, fewer than eight antenatal care contacts, maternal anaemia during pregnancy, and dependence on non-motorised transport when accessing health facilities (19, 20). Together, these factors point to the contribution of delayed engagement with maternity services, missed opportunities for early detection and management of pregnancy complications, and barriers to timely access to emergency obstetric care

## Conclusion

Overall, the findings indicate that implementation of the WHO Labour Care Guide over the eight-month post-implementation period was not associated with significant reductions in stillbirths, fresh stillbirths, birth asphyxia, or caesarean section rates. However, the absence of a large measurable effect in this study does not necessarily imply that the LCG is ineffective. Rather, it may reflect the influence of implementation-related factors, including the duration of follow-up, fidelity of use, provider adherence, documentation quality, and the readiness of emergency obstetric and newborn care systems. Longer follow-up, strengthened implementation support, and more detailed individual-level clinical data are needed to better determine the potential effect of the LCG on maternal and perinatal outcomes in routine maternity care settings.

## Strengths and Limitations

The study had several strengths. Firstly, it was guided by a clearly defined problem and objective, addressing an important maternal and newborn health priority in Zambia. The study design was appropriate because it enables comparison of outcomes before and after implementation of the WHO LCH across both implementation and comparison sites. The use of a DiD analytical approach further strengthened the evaluation by allowing assessment of changes over time while accounting for differences between study groups.

The inclusion of implementation and comparison sites improved the interpretation of findings and supported assessment of the effect of the intervention under routine service-delivery conditions. In addition, the target population and sample size were adequate for comparing outcomes and measuring implementation effects across the selected referral hospitals. Another important strength is that the study used real-world implementation data; therefore, the findings reflect actual clinical practice rather that controlled trial conditions. This enhances the practical relevance of the results for national scale-up, quality improvement, and policy decision-making.

On the other hand, the study had some limitations. The analysis relied on routinely collected HMIS/DHIS2 data, which limited the range and depth of variables available for analysis. As a result, the study could not comprehensively assess or compare some important maternal and newborn indicators, including duration of labour, obstructed labour, postpartum maternal complications, newborn survival beyond the immediate reporting period, and neonatal complications that may arise from suboptimal labour management, such as birth trauma. This limited the ability to explore the full pathway through which the WHO LCG may influence maternal and perinatal outcomes.

## Recommendations

Future evaluations of the WHO LCG should build on the strengths of this real-world implementation study by incorporating longer follow-up periods, stronger measurement of implementation fidelity, and more detailed individual-level clinical data. This would allow for a more comprehensive assessment of how the LCG influences labour progress, clinical decision-making, maternal complications, newborn outcomes, and longer-term neonatal survival.

Given that this study used routine HMIS/DHIS2 data, future studies should complement routine data with facility registers, patient records, LCG forms, and prospective clinical data collection to capture indicators not routinely reported, including duration of labour, obstructed labour, postpartum maternal complications, newborn resuscitation birth asphyxia, referral delays, and neonatal complication.

For national scale-up in Zambia, implementation of the LCG should be accompanied by sustained provider training, mentorship, supportive supervision, audit and feedback, and regular review of documentation quality. In addition, strengthening emergency obstetric and newborn systems, referral pathways, staffing, essential commodities, and decision-making capacity will be critical to maximizing the potential benefits of the LCG in routine maternal care settings.

## Data Availability

The data underlying the findings of this study are available from the corresponding author upon reasonable request, subject to applicable ethical and institutional requirements governing access to the study data.

## Declarations

### Ethical Oversight

This study was based on secondary routine health facility data. All data extracted from the District Health Information Management System 2 (DHIS2) were anonymised prior to analysis, and confidentiality was maintained throughout the study. Ethical approval was obtained from the University of Zambia Biomedical Research Ethics Committee (UNZABREC) (Ref. No. 7029-2025). Additional regulatory clearance was granted by the National Health Research Authority (NHRA) (Ref. No. NHRA-2672/08/09/2025).

### Consent for Publication

All authors have read and approved the final version of the manuscript.

### Availability of Data and Materials

The data and materials underlying this study are available from the Principal Investigator on reasonable request, subject to the necessary administrative and ethical clearance from the Copperbelt Provincial Health Office, Lusaka Provincial Health Office, and the Ministry of Health of the Government of the Republic of Zambia.

### Authors’ contributions

MM and BV contributed to the project inception and study conceptualization. MM led the study from inception and was responsible for the study design, data collection, data cleaning, data processing, data analysis, and manuscript drafting. MB and KL contributed to study conceptualization and manuscript review. BV provided oversight to ensure the quality, contextual relevance, and appropriate implementation of the research.

